# Radiation doses and Indications for Computed Tomography Scans among Paediatric Patients at a Tertiary Hospital in the Eastern Cape, South Africa

**DOI:** 10.64898/2026.03.21.26348958

**Authors:** Thembisa Mlamla, Anne Namugenyi, Juan Carlos Garcia-Alonso, Oladele Vincent Adeniyi

**Affiliations:** Department of Radiology, Nelson Mandela Academic Hospital, Mthatha, South Africa; Department of Family Medicine, Cecilia Makiwane Hospital, Mdantsane, East London, South Africa; Faculty of Health Sciences, Walter Sisulu University, Mthatha, South Africa

## Abstract

Computed Tomography (CT) provides superior diagnostic capability compared to plain radiography or ultrasound but involves higher ionising radiation doses, which is particularly concerning in children due to their increased radiosensitivity and longer life expectancy. In the absence of national CT diagnostic reference levels (DRLs) in South Africa, this study aimed to audit local DRLs at a tertiary hospital to establish the frequency and type of paediatric CT studies and benchmark institutional data against national and international standards. We conducted a retrospective cross-sectional study of 543 paediatric patients imaged between January 2021 and December 2023. Acquisition parameters, including CT Dose Index Volume (CTDIvol) and Dose Length Product (DLP) at the 75th percentile, were recorded for commonly imaged body regions. Results showed that the CT brain was the most frequent examination (86.6%). Mean CTDIvol values for the brain in age groups <1, 1–5, 6–10, and >10 years were 14.20, 14.20, 15.80, and 15.80 mGy, respectively, while mean DLPs were 343.70, 348.15, 415.18, and 485.53 mGy. Our findings indicate that institutional radiation doses are comparable to published national and international benchmarks. Notably, examinations conducted after hours exhibited slightly higher DRLs than those during normal working hours, suggesting a need for continued protocol optimization in all clinical shifts.

**Author Summary:** Medical imaging using CT is vital for diagnosing children, but it uses radiation that may increase long-term health risks due to their young age and sensitive bodies. In South Africa (SA), there are currently no national standards for how much radiation should be used for these scans. We conducted this study at a large hospital to audit our current practices and ensure we are keeping our youngest patients as safe as possible while still getting clear diagnostic images.

We looked at 543 CT scans performed on children over a 3 year period. We specifically looked at the radiation doses used for the most common scans, such as brain scans, across different age groups. Our results showed that the radiation levels at our hospital are in line with both international safety standards and locally. We also noticed that scans performed afterhours use slightly higher radiation doses than those during the day.

These results are encouraging because they show that our hospital is providing safe care that matches global benchmarks. However, the slightly higher doses during after-hours shifts suggest we should focus on more consistent training and standardized settings for all staff, regardless of when the scan is performed.

## Introduction

Computed tomography (CT) scan has become a cornerstone of modern medical diagnostic imaging (1). CT scans employ a cross-sectional imaging technique, which produces multiple slices of the body part and provides unparalleled diagnostic accuracy. CT studies are conducted quickly and rarely require additional resources, such as general anaesthesia, for paediatric patients. In comparison with magnetic resonance imaging (MRI), CT is cheaper and more accessible in low-to middle-income countries. This makes it the modality of choice in the initial assessment of critically ill and injured patients.

A major drawback with the use of CT scans is the ionising radiation they emit. CT scans have become the largest contributor to radiation exposure among patients (1). The radiation dose received is the most unwanted adverse effect of the CT scan, with the risk varying with age and the organ scanned (2). In children, exposure to high radiation doses may increase the risk of cancer and aplastic anaemia (3). The rapid cell division associated with growth in children makes their cells particularly susceptible to damage from ionising radiation (4). The life expectancy of children further increases their chances of developing cancerous transformation of the highly radiosensitive organs – breast, eyes, thyroid, colon and lungs (5). Previous studies by Mampuya et al. (6) and Fuji et al. (7) showed unequivocally that children tend to suffer more adverse effects from radiation than adults do.

The risk of malignant transformation is directly proportional to the radiation dose given. Therefore, ensuring shorter radiation times is an important goal in imaging. Where possible, alternative modalities such as MRI and ultrasound should be considered to reduce or avoid radiation exposure in children (8). However, MRI availability in resource-constrained settings remains a challenge. Paediatric-specific protocols play a vital role in reducing the dose of radiation in children (1). Among others, the campaign ‘Image Gently’ and the ‘as low as reasonably achievable’ (ALARA) principle are of critical importance in radiation protection among children (9, 10).

In order to maintain low radiation doses, the International Commission on Radiological Protection (ICRP) introduced the concept of diagnostic reference level (DRL) (11). DRLs are radiation dose reference levels set for common examinations to avoid unnecessary radiation. DRLs are expressed using many recognised dose metrics, but the most common include the volume-based CT dose index (CTDIvol) and the dose length product (DLP), among others (11, 12). CTDIvol indicates the average radiation dose delivered per slice, whereas the DLP represents the total radiation energy absorbed over the entire length of the scanned body part. This is calculated by multiplying the CTDIvol by the scan length (13). These measures enable more accurate monitoring of radiation exposure doses.

DRLs are useful in establishing standardised CT examinations across a region (14). In European and other developed countries, there are regulations requiring the establishment and maintenance of DRLs in departments where radiological equipment is used (15). In resource-limited countries, this practice has not been widely adopted, although they are the most constrained with equipment maintenance and upgrades (16). In South Africa, a few studies on DRLs for paediatric imaging (14, 16) have been undertaken, but many more are required to establish regional DRLs. Previous studies on paediatric CT DRLs from South Africa (17, 18) and Kenya (19) reported variations in paediatric DRLs. Given the lack of national protocols for paediatric CT scans in South Africa, it is imperative to audit local DLPs and CTDIvol and endeavour to standardise paediatric protocols for safe exposure to ionising radiation during CT examinations. The current study audited the radiation doses (CTDIvol and DLPs) used for paediatric CT examinations performed at a tertiary hospital in the Eastern Cape Province using the recommended age bands (20). In addition, the study examined variations in DRLs according to hours of work and anatomical sites imaged. Comparison was done with other studies done in the metropolitan cities in SA and internationally (14, 16, 18, 21)

The aim of the present study was to audit and describe local DRLs for paediatric CT scans at an academic tertiary referral centre in South Africa, and to compare them with data published in other tertiary hospitals in SA and globally. The study further examined variations in radiation dose by working hours and anatomical regions among paediatric patients.

## Methods

### Study design and setting

A retrospective cross-sectional study was conducted at the Radiology department of Nelson Mandela Academic Hospital (NMAH) in the Eastern Cape Province, South Africa. NMAH has a 700-bed capacity and serves as a tertiary referral centre for a population of about three million people. Approximately 7500 paediatric CT scans are performed annually.

### Study population and sample size estimation

Patients aged 0 to 14 years who had received CT scans during the period 1^st^ January 2021 to 31^st^ December 2023 were included in the study, irrespective of the time of the investigation, indication and the number of tests performed. Each CT scan was recorded as a separate event for analysis. Patients without CT scan images owing to cancellations were excluded from the study. The sample size was obtained by using Slovin’s formula:

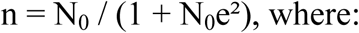

n is the sample size

N_0_ is the population size (7300)

e is the margin of error (0.05).

Given the retrospective nature of the study, an adjustment factor of 30% was applied to the total of 379 to achieve a sample of 493 CT scans, to account for anticipated missing values. Therefore, approximately 500 CT request forms and images stored on the PACS were retrieved and reviewed for this study. To ensure an equitable selection of CT scan images, a systematic sampling technique was adopted. A sampling interval of 15 was obtained with the formula N_0_/n (total estimated population divided by the sample size). The annual number of CT scan images (N_0_=7300) was divided by the sample size (n=500).

### Data collection

A self-designed proforma was used for data collection. Data was retrieved from the Picture Archiving and Communication System (PACS). The data were categorised according to the anatomical site and age group (18). A sampling interval of 15 was used, with every 15th CT scan image selected from the PACS.

### Measures

Relevant demographic information (age and sex) was obtained from the register at the CT scan unit. The age groups were divided into less than 1 year, 1–5 years, 6–10 years and > 10 years (18). In addition, indications for the CT scan and acquisition parameters were obtained from the PACS. Patient identity numbers were verified to avoid double-counting, except when different CT requests were made. Dates and times of the scans (whether conducted during or after working hours) were necessary to evaluate the effect of timing on the radiation dose, and were obtained from data captured on the CT scan. Anatomical region, i.e. head, chest, abdomen or neck, was recorded. The acquisition parameters CTDI_vol_ (measured in mGy) and DLP at the 75^th^ percentile (measured in mGycm) were recorded as displayed on the scan results.

The study used a Philips Ingenuity Core 128 slice scanner manufactured in Holland, The Netherlands. It is a multi-slice scanner, programmed in a helical mode, which incorporates pitch in the quantification of the dose output for the paediatric protocol. This was the reason why CTDIvol was the parameter considered instead of CTDIv. For each CT image, the date and time of the scan were recorded. Three different time categories were considered in the study: Monday to Friday from 08h00 to 16h00 (routine hours of work) and after hours from 16h00 to 8h00 the following day, and lastly, weekends and public holidays for 24h00. The categories were chosen as per recommendations made by the European Diagnostic Reference Levels for Paediatric Imaging Workshop in 2013 (20, 22).

### Statistical analysis

The statistical analysis was performed using the IBM SPSS Statistics for Windows, Version 27.0 (IBM Corp., Armonk, New York, USA). A summary of findings was generated using simple descriptive statistics. Radiation exposure levels (CTDI_vol_ and DLP dose indices) were categorised by patient age, time of scan, and anatomical site. For each age group and examination type, the median and 75th percentile (third quartile) values were calculated. Means with 95% confidence intervals were also reported for radiation dose levels. The results were compared with published South African DRLs and internationally recommended radiation dose levels. The 75th percentile values for CTDI_vol_ and DLP within each age and examination category were then compared with European and other national diagnostic reference levels (DRLs).

### Ethics approval

Ethical approval to conduct the study was obtained from the Ethics Committee of Walter Sisulu University (Reference: 137/2024). Permission was obtained from the Provincial Department of Health and the clinical governance of the participating tertiary hospital.

## Results

Overall, 543 CT scans were included in the analysis. The majority of the patients were male (59.5%). Nearly half (48.6%) were aged 6–10 years, followed by those older than 10 years (39.4%) (Table 1). Most CT examinations were performed during routine working hours (08h00–16h00), accounting for 76.4% of the scans.

**Table 1.**
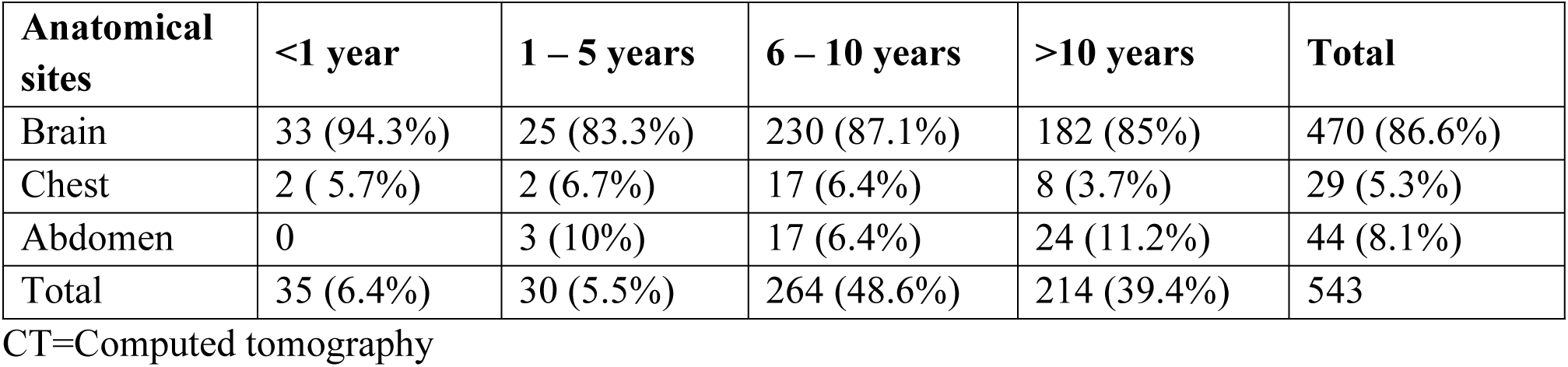
Disaggregation of CT scans by age and anatomical sites.

| Anatomical sites | <1 year | 1 – 5 years | 6 – 10 years | >10 years | Total |
| --- | --- | --- | --- | --- | --- |
| Brain | 33 (94.3%) | 25 (83.3%) | 230 (87.1%) | 182 (85%) | 470 (86.6%) |
| Chest | 2 ( 5.7%) | 2 (6.7%) | 17 (6.4%) | 8 (3.7%) | 29 (5.3%) |
| Abdomen | 0 | 3 (10%) | 17 (6.4%) | 24 (11.2%) | 44 (8.1%) |
| Total | 35 (6.4%) | 30 (5.5%) | 264 (48.6%) | 214 (39.4%) | 543 |

CT=Computed tomography

### Descriptive findings of the CT scans by age and anatomical sites

The most imaged body part was the brain (86.6%); this had a DLP value of 450.20 and a CTDIvol of 18.69. Abdomen CT scans were 44 (CTDIvol 9.19 and 343.44). Chest was the lowest with 29 (CTDIvol 10. 2 and DLP 388.31) CT studies (Table 2). Overall, there was a trend of general increase in radiation dose metrics (DLP and CTDIvol) with increasing age across brain, chest and abdominal CT examinations (Fig. 1).

**Fig. 1:**
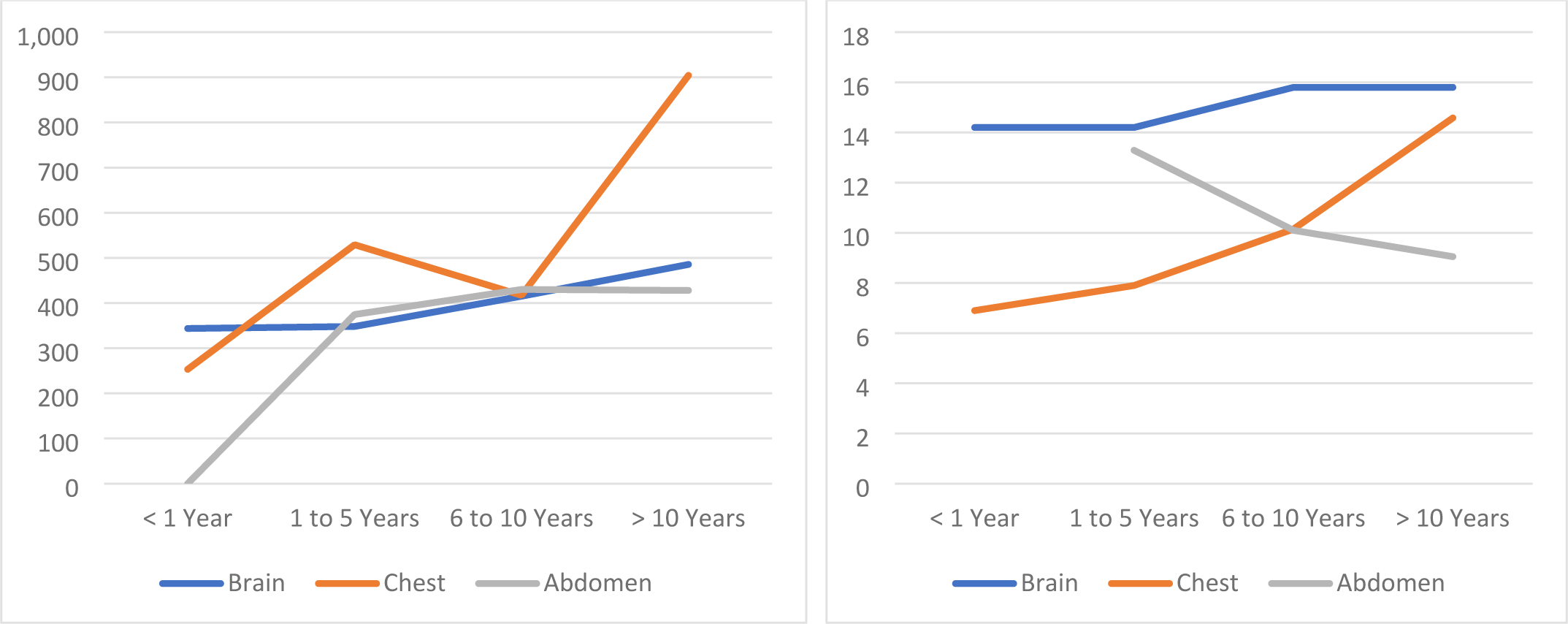
75^th^ percentile radiation dose metrics across paediatric age groups for brain, chest, and abdominal CT examinations; A=Dose length product B= CT dose index volume

**Table 2.**
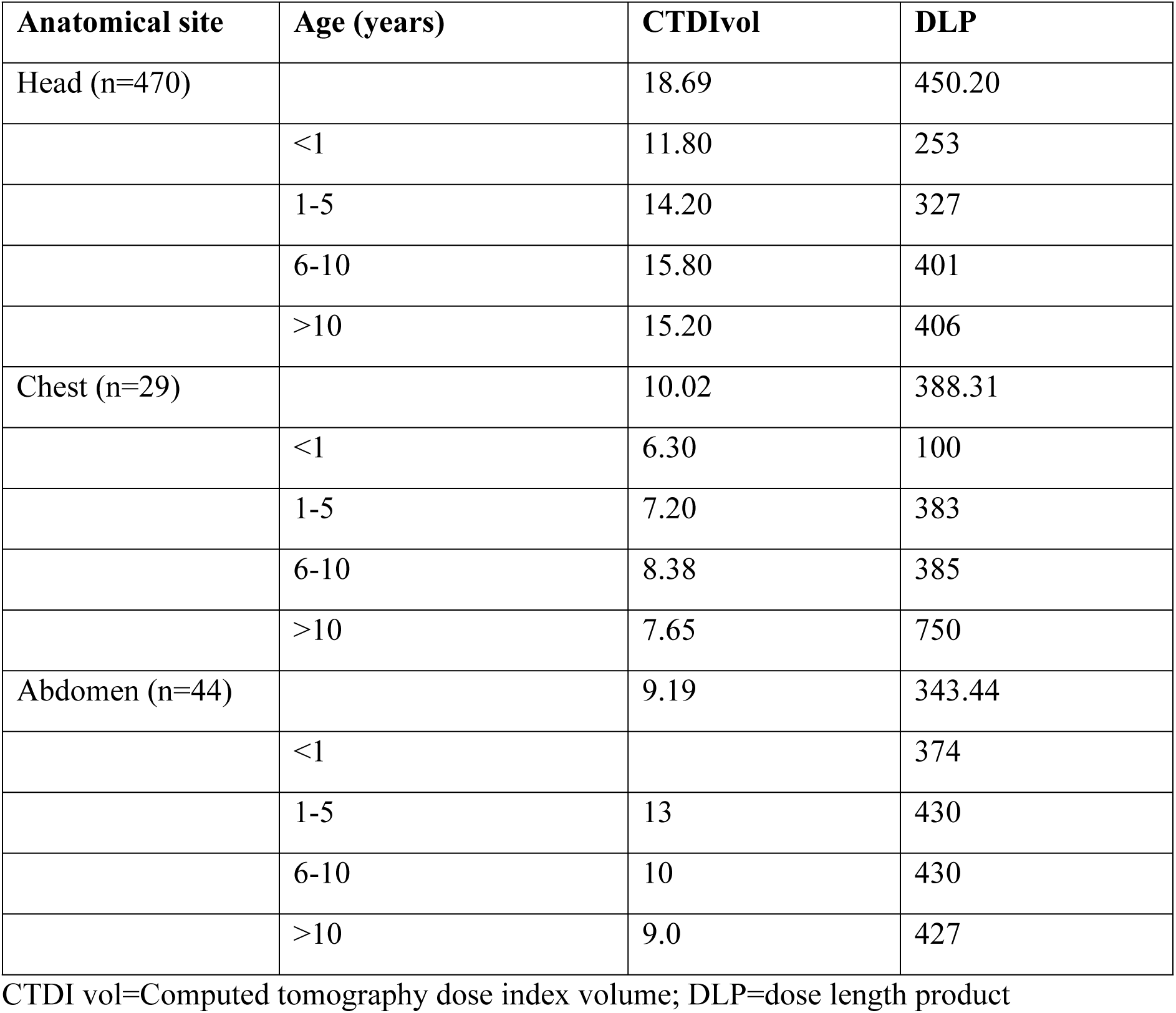
CTDI vol and DLP values by anatomical sites in different age groups.

| Anatomical site | Age (years) | CTDI<sub>vol</sub> | DLP |
| --- | --- | --- | --- |
| Head (n=470) |  | 18.69 | 450.20 |
|  | <1 | 11.80 | 253 |
|  | 1-5 | 14.20 | 327 |
|  | 6-10 | 15.80 | 401 |
|  | >10 | 15.20 | 406 |
| Chest (n=29) |  | 10.02 | 388.31 |
|  | <1 | 6.30 | 100 |
|  | 1-5 | 7.20 | 383 |
|  | 6-10 | 8.38 | 385 |
|  | >10 | 7.65 | 750 |
| Abdomen (n=44) |  | 9.19 | 343.44 |
|  | <1 |  | 374 |
|  | 1-5 | 13 | 430 |
|  | 6-10 | 10 | 430 |
|  | >10 | 9.0 | 427 |

CTDI vol=Computed tomography dose index volume; DLP=dose length product

### Descriptive findings of NMAH 75^th^ percentile CTDI_vol_

Overall, there was a slight variation in the 75^th^ percentile CTDI_vol_ according to age and time of day. For the under-one-year category, a lower mean CTDI_vol_ of 13.00 mGy during regular hours was recorded, compared to 15.00 mGy after hours during the week, and 13.80 mGy on weekends. However, there was no variation in the CTDI_vol_ by time of day for children 1–5 years (14.20 mGy) and 6–10 years (15.80 mGy). A slight variation in CTDI_vol_ was, however, observed among children over 10 years (19.82 mGy) during regular hours, compared to 15.80 mGy after-hours and weekends (Table 3).

**Table 3.** Descriptive findings of NMAH 75th percentile CTDI_vol_ values in mGy.

| **Age** | **Regular hours** | **After-hours** | **Weekends** | **Combined** |
| --- | --- | --- | --- | --- |
| <1 | 13.00 | 15.00 | 13.80 | 14.20 |
| 1 – 5 | 14.20 | 14.20 | 14.20 | 14.20 |
| 6 – 10 | 15.80 | 15.80 | 15.80 | 15.80 |
| >10 | 19.92 | 15.80 | 15.80 | 15.80 |

CTDI vol=Computed tomography dose index volume; NMAH=Nelson Mandela Academic Hospital

### Descriptive finding of 75^th^ percentile of CTDIvol, DLP and effective dose

The CTDIvol for the brain remained consistent across all age groups, with values of approximately 14-16 mGy, although the DLP increased with age, reflecting longer scan lengths in older children. The number of contributing studies for chest CT was relatively small, especially among the younger age group, limiting the reliability of the estimates. Abdominal CT data were unavailable for children younger than 1 year (Table 4)

**Table 4:** 75^th^ percentile of Computed Tomography Dose Index Vol (mGy), Dose Length Product (mGy·cm), and Effective Dose (mSv) for each computed tomography anatomical site, in each age group, as well as the total number of contributing studies per category (n = 543).

| Anatomical site | Age group (years) | CTDI<sub>Vol</sub>: 75<sup>th</sup> percentile |  | DLP: 75<sup>th</sup> percentile |  | Effective dose 75<sup>th</sup> percentile |  | No of studies |
| --- | --- | --- | --- | --- | --- | --- | --- | --- |
|  |  | 95% | CI | 95% | CI | 95% | CI |  |
| Head | <1 | 14.20 | 11.80 – 26.25 | 343.70 | 253.21 – 715.80 | 2.62 | 2.03 – 7.83 | 33 |
|  | 1 – 5 | 14.20 | 14.20 – 14.20 | 348.15 | 327.70 – 388.58 | 2.26 | 2.13 – 2.58 | 25 |
|  | 6 – 10 | 15.80 | 15.80 – 15.80 | 415.18 | 401.00 – 437.00 | 1.39 | 1.30 – 1.47 | 230 |
|  | >10 | 15.80 | 15.80 – 30.92 | 485.53 | 406.91 – 605.00 | 1.55 | 1.30 – 1.94 | 182 |
| Chest | <1 | 6.90 | 6.30 – 6.90 | 253.20 | 100.70 – 253.20 | 6.58 | 2.62 – 6.58 | 2 |
|  | 1 – 5 | 7.90 | 7.20 – 7.90 | 529.20 | 383.80 – 529.20 | 13.76 | 9.98 – 13.76 | 2 |
|  | 6 – 10 | 10.15 | 8.38 – 5.8 | 417.15 | 385.60 – 805.60 | 5.42 | 5.01 – 10.47 | 17 |
|  | >10 | 14.58 | 7.65 – 63.30 | 904.33 | 750.10 – 1457.60 | 11.76 | 3.51 – 18.95 | 8 |
| Abdomen | <1 | – | – | – | – | – | – | 0 |
|  | 1 – 5 | 13.29 | 10.00 – 13.48 | 374.64 | 284.79 – 446.80 | 11.24 | 7.21 – 13.40 | 3 |
|  | 6 – 10 | 10.10 | 7.60 – 15.80 | 430.15 | 335.45 – 503.80 | 6.45 | 5.03 – 7.56 | 17 |
|  | >10 | 9.05 | 8.90 – 14.20 | 427.98 | 381.10 – 479.30 | 6.42 | 5.72 – 7.19 | 24 |

CTDI vol=Computed tomography dose index volume; CI=Confidence interval; DLP=dose length product;

## Discussion

### Patterns of Paediatric CT examinations

A substantial majority of the children in this study were over the age of five years (88%), with slightly more boys accessing CT scans, which corroborates what was documented by Van der Merwe et al (18). Children in this age category are more prone to injuries, among other conditions, necessitating imaging for traumatic brain injury (26). This observation is supported by the study finding that the substantial majority of images were obtained for brain CT (86.6%). While magnetic resonance imaging (MRI) of the brain may be the ideal modality, it is not easily accessible and most times requires anaesthetic services. There were fewer chest and abdominal CT examinations, just like what was reported in another study (18). Very few chest and abdomen CT studies were recorded among the infants and toddlers in the study period. This is an important finding showing that the institution abides by the imaging gently principle (10). Ultrasound is the modality of choice in this age group, and where results are inconclusive, these patients are prioritised for MRI.

The audit established that CT brain was the most commonly imaged paediatric study (86.6%) with 75^th^ percentile CTDIvol and DLP values that are favourably comparable to those published by other studies locally and internationally (14, 18, 21). This excellent agreement of these baseline DRLs at our institution suggests that CT brain protocols and technical parameters meet international standards and are similar to other tertiary hospitals within the country (14, 18, 23).

It was observed that the number of chest and abdomen CT scans among the age group of the less than 5-year-old were not statistically significant for analysis. The ICRP/DRL survey recommends at least 30 standardised studies to be included when establishing local DRLs (14, 20). An objective comparison, therefore, could not be done for CT chest and abdomen in these two age groups in the current study. However, among the 6-10 and over 10 years categories, the mean DLPs levels were higher than the 75^th^ percentile by over 100% compared to registered national and international levels (14, 21). This reflects suboptimal scanning techniques, highlighting a need for urgent review of the chest and abdominal scanning protocols at the institution. One major contributing factor for the high radiation doses, is the use of multiple phase protocols rather than the single phase used in European countries (27). In a resource-constrained environment where it is not possible to review studies before patients get off the CT table, multiphase protocols are used to reduce repeat CTs and misses (12).

### Trends of Radiation Doses

There was no significant percentage increment in radiation doses recorded after hours and over the weekends and public holidays. While specific acceptable percentage variations for DRLs are not universally defined, some studies report a variation of 20-80% as being significant and must trigger protocol review (28). A linear relationship between radiation doses (both DLPs and CTDI_vol_) and the ages of the patients was noted. Smaller doses were documented in children under one year, and higher doses among the older children. A similar trend of increasing radiation doses with increasing age was observed for CT scans of both the chest and abdomen. This observation of a linear relationship between CTDIvol and DLP across age groups suggests consistent protocol application and appropriate scaling of radiation dose with patient size (29). This supports the reliability of dose data and indicates that dose optimisation practices were generally maintained.

### Comparison with National and international DRLs

DRLs for CT brain among children for all age groups were found to be within the acceptable 10 to 20% increase compared to the established DRLs nationally and other regions (Table 5). The audit established that CT brain DLP values recorded at NMAH for all age groups ranged from 343.7 to 485.53 mGy.cm. These values are comparable to the Johannesburg hospital DLPs (311.3 - 746.1mGy.cm) and the European (300 - 650 mGy.cm) benchmark values. When compared to similar middle to lower-income countries (Kenya and Brazil), their DLPs were much higher (Table 5). This observation, where the institutional baseline DRLs were comparable to local and international DRLs, attests to the use of the right protocols in the most common paediatric CT study.

**Table 5:** Dose Length Product 75th percentile (mGy·cm) of NMAH hospital compared to Johannesburg hospitals and international diagnostic reference levels.

| **Examination Grouped in years** | **NMAH** | **No NMAH** | **Jhb\*** | **No Jhb** | **EDRL†** | **Germany§** | **Japan¶** | **Kenya††** | **Brazil‡‡** |
| --- | --- | --- | --- | --- | --- | --- | --- | --- | --- |
| **Brain** |  |  |  |  |  |  |  |  |  |
| <1 | 343.70 | 33 | 311.30 | 169 | 300 | 300 | 50 | 1005 | 290 |
| 1–5 | 348.15 | 25 | 362.40 | 238 | 385 | 450 | 660 | 1395 | 550 |
| 6–10 | 415.18 | 230 | 457.20 | 156 | 505 | 650 | 850 | 1608 | 670 |
| >10 | 485.53 | 182 | 746.10 | 124 | 650 | 800 | – | – | 880 |
| **Chest** |  |  |  |  |  |  |  |  |  |
| <1 | 253.20 | 2 | 153.50 | 12 | 35–50 | 25 | 210 | – | 64 |
| 1–5 | 529.20 | 2 | 105.10 | 9 | 50–70 | 55 | 300 | 215 | 130 |
| 6–10 | 417.15 | 17 | 136.40 | 8 | 70–115 | 110 | 410 | – | – |
| >10 | 904.33 | 8 | 325.10 | 7 | 115–200 | 200 | – | 453 | – |
| **Abdomen** |  |  |  |  |  |  |  |  |  |
| <1 | – | 0 | 191.50 | 4 | 45–120 | – | 220 | – | 110 |
| 1–5 | 374.64 | 3 | 187.80 | 18 | 120–150 | – | 400 | 764 | 170 |
| 6–10 | 430.15 | 17 | 203.30 | 14 | 150–210 | 185 | 530 | – | 220 |
| >10 | 427.98 | 24 | 371.10 | 26 | 210–480 | 310 | – | – | – |

The above table was adopted from (18): \* Johannesburg(18); †European Commission (2018) Radiation Protection No 185 (14); ‡Doses from Computed Tomography (CT) Examinations in the UK-2011 (15); §Bundesamt für Strahlenschutz (2016) (21); ¶Japan Network for research and Information on Medical exposures (2015) (23); ††National Diagnostic Reference Level Initiative for Computed Tomography examinations in Kenya (2016) (24); ‡‡A Contribution to the Establishment of Diagnostic Reference Levels in Computed Tomography in Brazil (25)

High values were documented among the chest CT studies in the age group of 5-10 and 10-14 year-olds, with a percentage increase of 179% compared to the established international DRLs. The European guideline does not have strict levels or cut-off of DRLs, however variations of such magnitude have triggered protocol review in the institution. For chest CT examination in the infants and toddlers (<1 to 5 years), the number of studies was way below the recommended European guideline of at least 20 patient examinations per protocol to determine local DRLs (14).

There were no abdominal CT examinations among children below 1 year and very few were recorded among the 1 to 5 year olds. In the 6-year old and above, CT abdomen DRLs at NMAH were higher than those at the Johannesburg hospitals by more than 100%. Higher radiation doses for chest and abdominal CT scans in the current study could possibly be due to the high burden of infectious diseases, notably tuberculosis, in the region (30).

### Study limitations

This was a single-centre study with relatively limited data, especially for the under five-year-olds. Also, there were a few chest and abdominal CT scans, making it impossible to generalise the findings. Nonetheless, this study provides baseline data for the Eastern Cape tertiary academic centres for reference and highlights critical issues for urgent attention.

### Conclusions

The study established local DRLs for the institution for the most common paediatric CT studies. It was satisfying to note that for the most common CT studies, the institution is at par with national and international DRLs. Diagnostic reference levels (DLPs and CTDIvol values) for procedures are important tools for optimising patient radiation doses and for identifying instances where these levels may be unusually high. This is particularly relevant for paediatric patients undergoing CT due to the associated relatively high radiation exposure.

## Contribution

The outcome of this study was the increased awareness among CT staff regarding the radiation doses delivered to the paediatric patients. The radiation doses for most CT examinations across all age groups were comparable with both national and international benchmarks. Given that appropriate protocol optimisation processes were followed is very reassuring. However, for chest and abdominal CT examinations, the institutional DRLs were higher than the benchmark values, highlighting the need for urgent protocol review, optimisation and justification.

## Data Availability

n/a

## Acknowledgement

The authors are grateful to Dr Ncube for the support and guidance during the implementation of the study.

## Abbreviations

ALARA: As Low As Reasonably Achievable
CT: Computed Tomography
CTDI100: Computed Tomography Dose Index over 100 mm.
CTDIw: Computed Tomography Dose Index weighted for centre and periphery of phantom
CTDIvol: Computed Tomography Dose Index volume
DLP: Dose Length Product
DRL: Diagnostic Reference Level
ICRP: International Commision on Radiological Protection mGy milligray mGy*cm milligray-centimetre
MRI: Magnetic Resonance Imaging
PACS: Picture Archive and Communications System

## Notes

### Competing Interest Statement

The authors have declared no competing interest.

### Funding Statement

The author(s) received no specific funding for this work.

